# Panbio antigen rapid test is reliable to diagnose SARS-CoV-2 infection in the first 7 days after the onset of symptoms

**DOI:** 10.1101/2020.09.20.20198192

**Authors:** Manuel Linares, Ramón Pérez-Tanoira, Juan Romanyk, Felipe Pérez-García, Peña Gómez-Herruz, Teresa Arroyo, Juan Cuadros

## Abstract

**Background:** Real-time reverse transcription polymerase chain reaction (RT-qPCR) is the current recommended laboratory method to diagnose SARS-CoV-2 acute infection, several factors such as requirement of special equipment, time consuming, high cost and skilled staff limit the use of these molecular techniques. A more rapid and high-throughput method is in growing demand.

**Methods:** Evaluate the performances of the Panbio™ COVID-19 AG Rapid Test Device (Nasopharyngeal), a rapid immunochromatographic test for the detection of SARS-CoV-2 antigen, in comparison to RT-qPCR.

**Results:** The RDT evaluated in this study showed a high sensitivity and specificity in samples mainly obtained during the first week of symptoms and with high viral loads.

**Conclusions:** This assay seems to be an effective strategy for controlling the COVID-19 pandemic for the rapid identification and isolation of SARS-CoV-2 infected patients.

## Introduction

Ensuring accurate diagnosis is essential to limit the spread of SARS-CoV-2 and for the clinical management of COVID-19. Although real-time reverse transcription polymerase chain reaction (RT-qPCR) is the current recommended laboratory method to diagnose SARS-CoV-2 acute infection, several factors such as requirement of special equipment, time consuming, high cost and skilled staff limit the use of these molecular techniques. A more rapid and high-throughput method is in growing demand. (1,2)

Until now, the use of antigen detection tests alone had been ruled out and not recommended due to their low sensitivity (3-5). Previously in the first wave, several easy to perform rapid antigen detection tests were developed as the first line of diagnostic. However, the results obtained were not good enough. (6-9)

## Material and methods

### Population and study period

Study was conducted between September 10, 2020 and September 15, 2020. We included patients that were attended at our emergency department and in two of our primary healthcare centers:

#### Emergency department

(ED) patients: we included 135 symptomatic patients that were admitted in our ED with clinical suspicion or COVID-19 and 17 asymptomatic patients with history of contact with another COVID-19 patient.

#### Primary healthcare

(PH) patients: 50 symptomatic patients and 55 asymptomatic patients attendend in two of our primary healthcare centres.

A total of two consecutive nasopharyngeal swabs were obtained from each patient. One of them was employed to perform the antigenic rapid test and the other sample was employed to carry out the RT-PCR.

### Diagnostic procedures

#### RT-PCR

one automatic extractor was employed to obtain viral RNA from clinical samples: Hamilton Microlab Starlet (Hamilton Company, Bonaduz, Switzerland). RNA amplification was made using Allplex SARS-CoV-2 assay (Seegene, Seoul, South Korea).

#### Antigenic rapid test

we applied the Panbio COVID-19 Ag Rapid Test Device (Abbott Rapid Diagnostic Jena GmbH, Jena, Germany). This test is a qualitative membrane-based immunoassay (immunochromatography) for the detection of Nucleocapsid protein of SARS-CoV-2 in nasopharyngeal samples.

### Clinical data

Demographic (age and sex) and clinical variables of the study population were obtained from the medical records. We also recorded the time from the onset of symptoms and the story of prior contact with COVID-19 patients.

### Statistical analysis

Specificity and sensitivity with 95% confidence intervals (95%CI) were calculated using the RT-PCR results as gold standard. Sensitivity was evaluated globally and also according to the time from the onset of symptoms (< 7 days and > 7 days). Additionally, we also calculated the negative predictive value (NPV) and positive predictive value (PPV) for estimated prevalence of infection of 1%, 2.5%, 5% and 10%. Agreement between techniques was evaluated using Cohen’s kappa score. (10)

Continuous variables were presented as median and interquartile range (IQR) and categorical variables as proportions. We used the Mann-Whitney U-test, χ2 test, or Fisher’s exact test to compare differences between survivors and non-survivors where appropriate. For these comparisons, a p value of 0.05 or below was considered significant. Statistical analysis was performed with SPSS v20.0 (IBM Corp., Armonk, NY, USA).

## Results

255 nasopharyngeal swabs collected from symptomatic and asymptomatic patients were tested. 60 (23,5%) positive RT-qPCR samples, 44 (17,2%) were detected by the rapid antigen test. Overall, amongst the 60 positive samples, the Ct value was detected for gene N in 53 samples. The COVID-19 Ag detected 43 samples (81.1%) (Table 3). The range of cycle threshold (Ct) values was 10.84-37.65 (median 22.32 IQR 18.31-29.02). For samples with Ct < 25 (n = 33), <30 (n = 7), <35 (n = 2) and <40 (n=1), COVID-19 Ag has a sensitivity of 100%, 87.5%, 25% and 25% respectively. However, in our study population, the overall sensitivity is 73.3% (95% IC: 62.2–83.8). Considering only symptomatic patients with less of seven days since onset, the sensitivity is 86.5% (95% IC: 75.0-97.0). Specificity is always 100%.

**Table 1.**
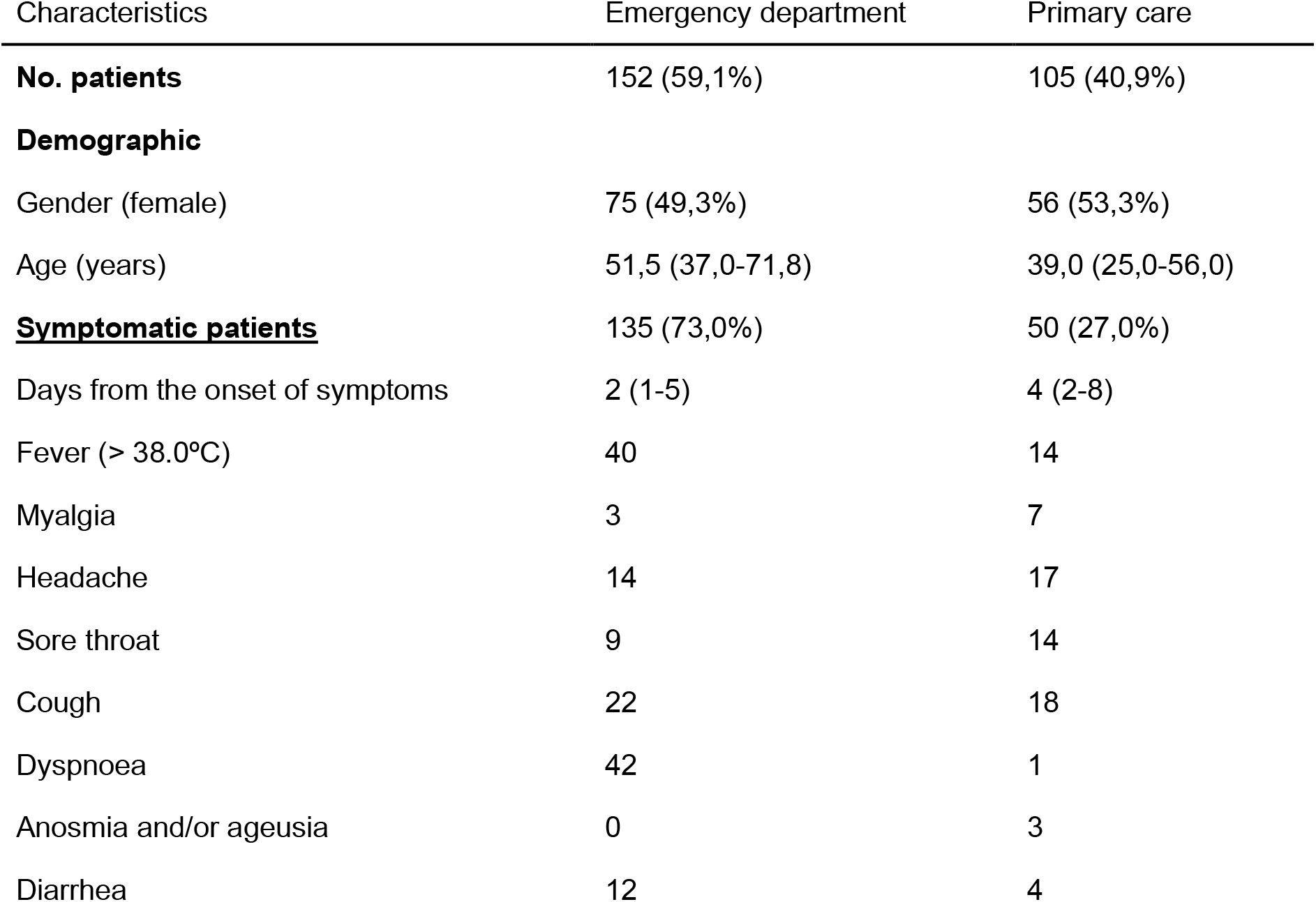
Patients.

**Table 2.**
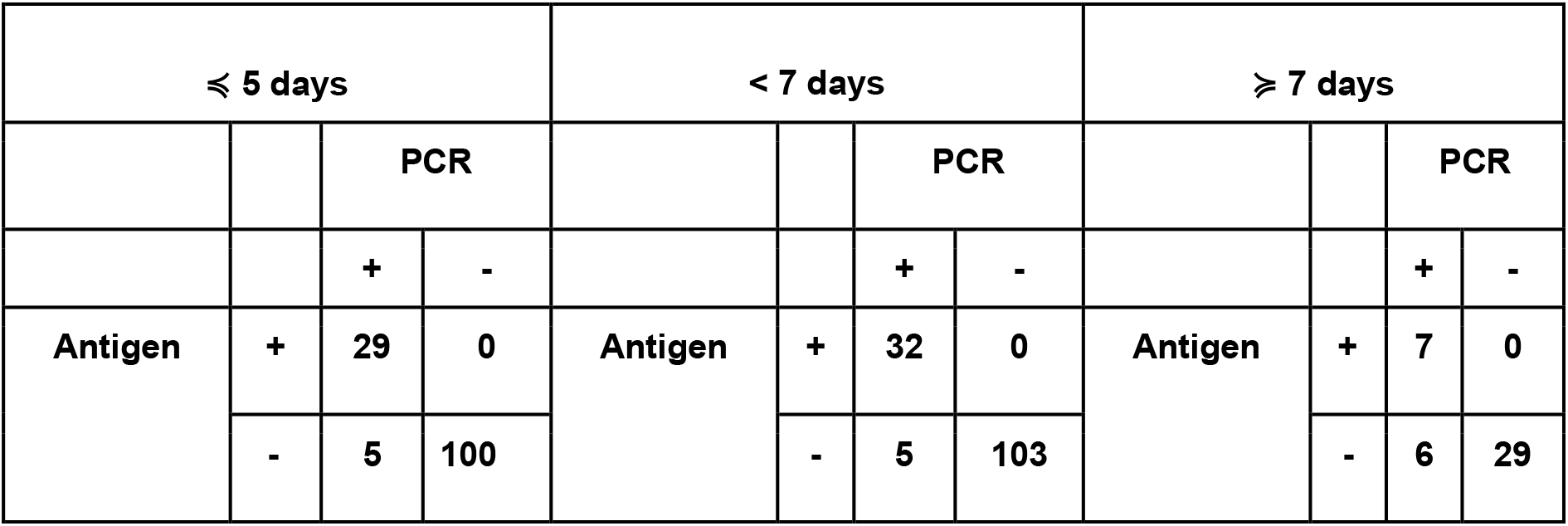
Agreement between antigenic test and PCR.

**Table 3.**
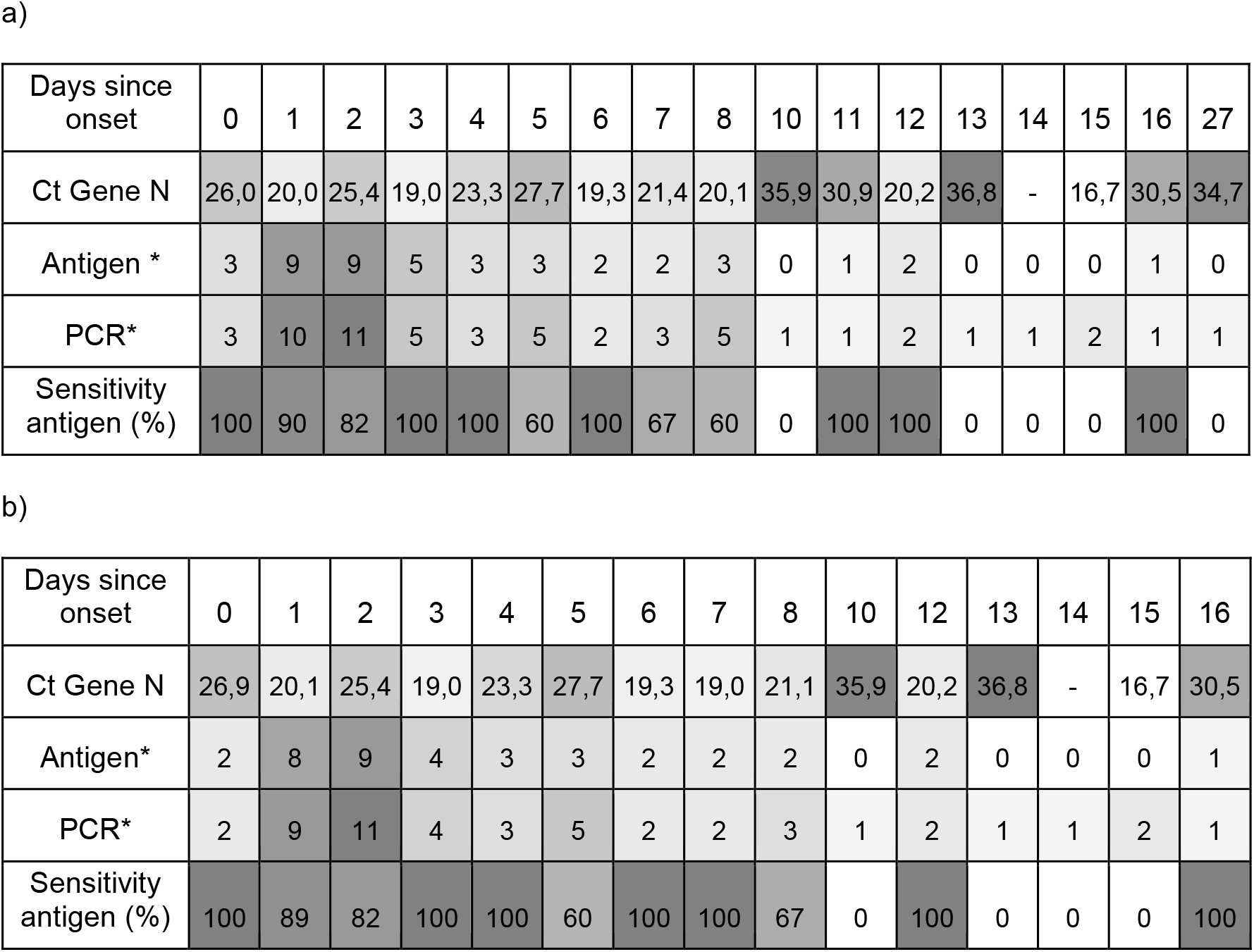
Color scale stepped from white to gray of median of Cycle threshold (Ct), number of positive results for antigen test and PCR test and sensitivity of antigen test considering days since onset of **a)** symptoms or contact and **b)** only symptoms. *Number of positive results

The RDT evaluated in this study showed a high sensitivity and specificity in samples mainly obtained during the first week of symptoms and with high viral loads (table 2)

**Figure 1.**
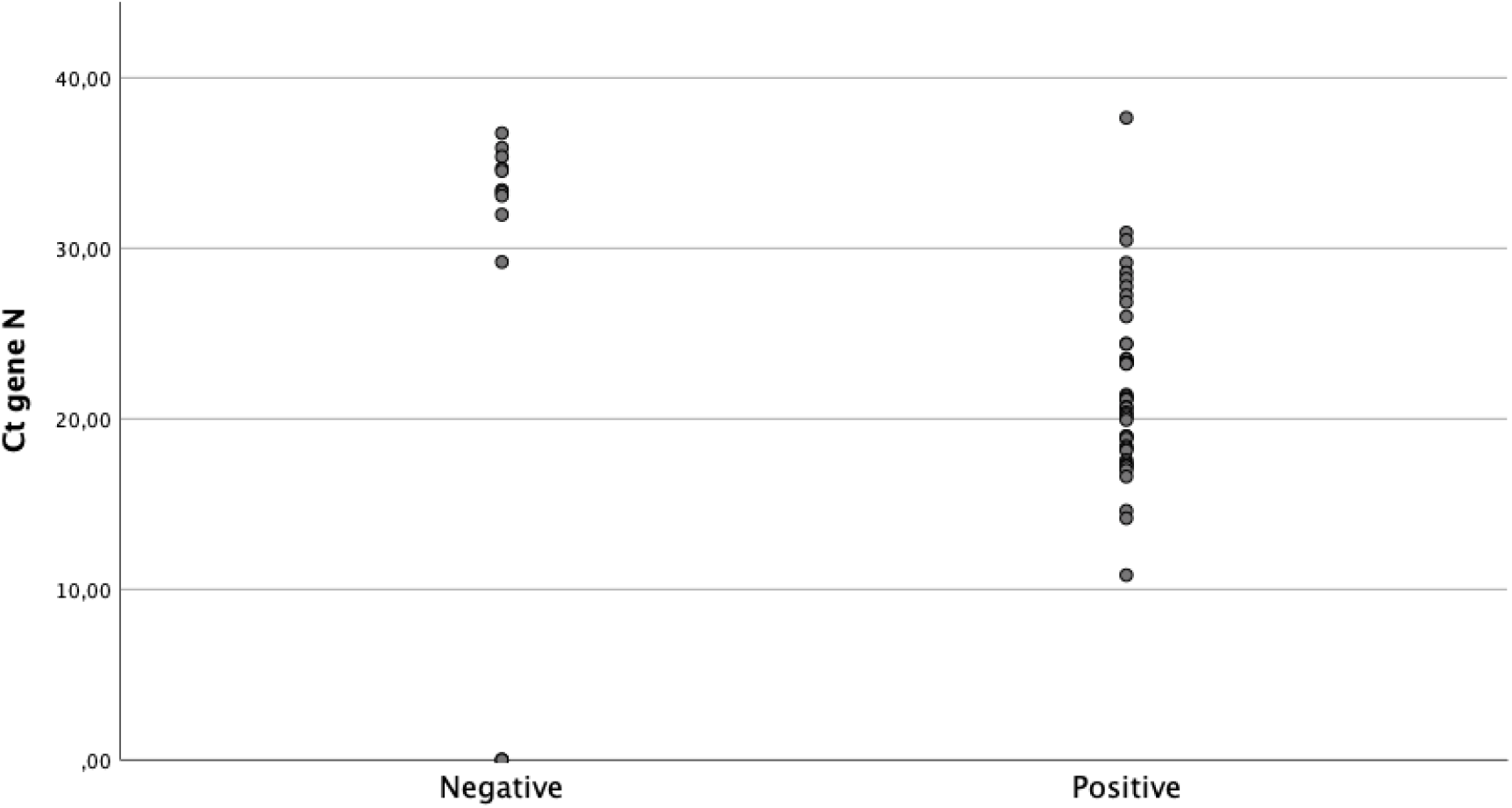
COVID-19 antigen results according to viral load.

## Discussion

As of September 18, 2020, the Food and Drug Administration (FDA) has approved more tan 150 SARS-CoV-2 RNA detection kits. (11,12) The lateral flow assay (LFA) is user-friendly, cheap, and easily mass-produced. In addition, it may be the optimal method for in-field detection of SARS-CoV-2 antigens if accurate, easy-to-use, rapid, and cost-effective. (13)

Our results suggest that Panbio ™ COVID-19 AG Rapid Test Device can rapidly identify SARS-CoV-2-infected individuals with moderate to high viral loads. Antigenic tests have shown 100% specificity in all types of patients and can be a powerful tool of high positive predictive value to control the COVID pandemia. It could be used as a substitute technique for PCR in this type of patients to shortcut delays and intensive labour costs generated by the massive use of PCRs.

In the group of symptomatic patients with less than 7 days of evolution, the test reaches a sensitivity of 86,5%, low to that described in the technical data sheet of the test. It is necessary to carry out studies with more patients to assess the usefulness that this technique could have in evaluating the infective potential of close contacts with PCR positive patients (14,15). When testing with antigen tests, it must be considered that the infection prevalence and the clinical context of the recipient of the test affects at the pretest probability of the result being correct. Also, rapid antigen tests could be used for screening in high-risk clusters settings to identify quickly persons with a SARS-CoV-2 infection and prevent the transmission by repeat testing.

## Data Availability

The data that support the findings of this study are available from the corresponding author, M.Linares, upon reasonable request.

## Abbreviations

COVID-19: Coronavirus disease 2019
SARS-CoV-2: Severe acute respiratory syndrome coronavirus 2
NP: Nucleocapsid protein
LFA: Lateral flow assay
FDA: Food and Drug Administration
RT-qPCR: Real-time reverse transcription polymerase chain reaction
RDT: Rapid diagnostic test.

## References

1. Loeffelholz MJ, Tang YW. Laboratory diagnosis of emerging human coronavirus infections - the state of the art. Emerg Microbes Infect. 2020;9(1):747–756. doi:10.1080/22221751.2020.1745095

2. Lieberman JA, Pepper G, Naccache SN, Huang ML, Jerome KR, Greninger AL. Comparison of Commercially Available and Laboratory-Developed Assays for In Vitro Detection of SARS-CoV-2 in Clinical Laboratories. J Clin Microbiol. 2020;58(8):e00821–20. Published 2020 Jul 23. doi:10.1128/JCM.00821-20

3. Coronavirus (COVID-19) Update: FDA Authorizes First Antigen Test to Help in the Rapid Detection of the Virus that Causes COVID-19 in Patients (Stephen M, Hahn M.D. 2020 May 09: Commisioner of Food and Drugs

4. Scohy A, Anantharajah A, Bodéus M, Kabamba-Mukadi B, Verroken A, Rodriguez-Villalobos H. Low performance of rapid antigen detection test as frontline testing for COVID-19 diagnosis. J Clin Virol. 2020;129:104455. doi:10.1016/j.jcv.2020.104455

5. Nagura-Ikeda M, Imai K, Tabata S, et al. Clinical Evaluation of Self-Collected Saliva by Quantitative Reverse Transcription-PCR (RT-qPCR), Direct RT-qPCR, Reverse Transcription-Loop-Mediated Isothermal Amplification, and a Rapid Antigen Test To Diagnose COVID-19. J Clin Microbiol. 2020;58(9):e01438–20. Published 2020 Aug 24. doi:10.1128/JCM.01438-20

6. In Vitro Diagnostic Assays for COVID-19: Recent Advances and Emerging Trends (Sandeep Kumar Vashist, 2020 April 05: diagnostics)

7. D. Das, S. Kammila, M.R. Suresh. Development, characterization, and application of monoclonal antibodies against severe acute respiratory syndrome coronavirus nucleocapsid protein Clin. Vaccine Immunol.: CVI, 17 (12) (2010), pp. 2033–2036

8. Kyosei Y, Namba M, Yamura S, et al. Proposal of De Novo Antigen Test for COVID-19: Ultrasensitive Detection of Spike Proteins of SARS-CoV-2. Diagnostics (Basel). 2020;10(8):E594. Published 2020 Aug 14. doi:10.3390/diagnostics10080594

9. Mak GC, Cheng PK, Lau SS, et al. Evaluation of rapid antigen test for detection of SARS-CoV-2 virus. J Clin Virol. 2020;129:104500. doi:10.1016/j.jcv.2020.104500

10. M.L. McHugh, Interrater reliability: the kappa statistic, Biochem Med (Zagreb). 22 (2012) (276–282).

11. FDA. Coronavirus Disease 2019 (COVID-19) Emergency Use Authorizations for Medical Devices. Retrieved Sep 18, 2020. https://www.fda.gov/medical-devices/emergency-useauthorizations-medical-devices/coronavirus-disease-2019-covid-19-emergency-useauthorizations-medical-devices

12. Information for laboratories about coronavirus (COVID-19). (2020). Retrieved Sep 18, 2020, from https://www.cdc.gov/coronavirus/2019-ncov/lab/resources/antigen-testsguidelines.html

13. Dinnes J, Deeks JJ, Adriano A, et al. Rapid, point-of-care antigen and molecular-based tests for diagnosis of SARS-CoV-2 infection. Cochrane Database Syst Rev. 2020;8:CD013705. Published 2020 Aug 26. doi:10.1002/14651858.CD013705

14. CDC. Discontinuation of Transmission-Based Precautions and Disposition of Patients with COVID-19 in Healthcare Settings (Interim Guidance). (2020). Retrieved Sep 18, 2020 https://www.cdc.gov/coronavirus/2019-ncov/hcp/disposition-hospitalized-patients.html

15. CDC. Duration of Isolation and Precautions for Adults with COVID-19. (2020). Retrieved Sep 18, 2020 https://www.cdc.gov/coronavirus/2019-ncov/hcp/durationisolation.html#:~:text=Recommendations,-Duration%20of%20isolation&text=For%20most%20persons%20with%20COVID,with%20improvement%20of%20other%20symptoms.

